# Carotid body dysregulation contributes to the enigma of long COVID

**DOI:** 10.1101/2023.05.25.23290513

**Authors:** Ahmed El-Medany, Zoe H Adams, Hazel C Blythe, Katrina A Hope, Adrian H Kendrick, Ana Paula Abdala Sheikh, Julian FR Paton, Angus K Nightingale, Emma C Hart

## Abstract

The symptoms of long COVID, which include fatigue, breathlessness, dysregulated breathing, and exercise intolerance, have unknown mechanisms. These symptoms are also observed in heart failure and are partially driven by increased sensitivity of the carotid chemoreflex. As the carotid body has an abundance of ACE2 (the cell entry mechanism for SARS-CoV-2), we investigated whether carotid chemoreflex sensitivity was elevated in participants with long COVID. During cardiopulmonary exercise testing, the V_E_/VCO_2_ slope (a measure of breathing efficiency) was higher in the long COVID group than in the controls, indicating excessive hyperventilation. The hypoxic ventilatory response, which measures carotid chemoreflex sensitivity, was increased in long COVID participants and correlated with the V_E_/VCO_2_ slope, suggesting that excessive hyperventilation may be related to carotid body hypersensitivity. Therefore, the carotid chemoreflex is sensitized in long COVID and may explain dysregulated breathing and exercise intolerance in these participants. Tempering carotid body excitability may be a viable treatment option for long COVID patients.

## Introduction

Long COVID (post-COVID-19 syndrome), is a multi-organ, often debilitating condition associated with a range of symptoms. The UK’s National Institute for Health Care and Excellence (NICE) defines long COVID as ongoing symptoms lasting for 12 or more weeks after initial SARS-CoV-2 infection, without alternative explanations^1^. The estimated incidence of long COVID varies, and is reported to be up to 41% of non-hospitalised cases ^2 3 4^ and up to 76% of hospitalised cases^5 6^. The prevalence decreases in vaccinated populations^7^. In January 2023, 2 million people self-reported long COVID symptoms in the UK, with 77% experiencing adverse effects in their daily activities^8^. Persistent symptoms include chronic fatigue, ‘brain fog,’ cognitive impairment and memory loss, dyspnoea at rest and on exertion, exercise intolerance, orthostatic intolerance, inappropriate postural tachycardia, and episodic hyperadrenergic surges^4,9,10^. A meta-analysis of 63 studies worldwide, with a total COVID-19 population of 257,348, reported that between 3-6 and 9-12 months post-infection, fatigue and dyspnoea were the most reported symptoms, with a prevalence of 32-47% and 21-25% respectively^11^. Despite the prevalence of long COVID and severely disabling symptoms there are no treatment strategies available. Thus, it is crucial to identify the mechanisms involved in long COVID to inform urgently needed therapy. It is likely that mechanisms depend on the severity of the original infection and are different for hospitalised (e.g. long term sequalae from intensive care and intubation/ventilation) versus non-hospitalised patients who had mild to moderate initial symptoms.

Currently, the exact mechanisms driving long COVID in non-hospitalised patients remain unknown but are likely to be multiple^12,13^. Studies have shown that exercise intolerance and disorganised breathing or breathing inefficiency during exercise are key features of long COVID ^14–17^ even in patients who have normal lung function and no evidence of gas exchange abnormalities^16^. The carotid bodies are key oxygen, carbon dioxide and pH sensing organs that control ventilation, dyspnoea, and the circulation at rest and during exercise in health and disease^18,19^. In fact, in chronic heart failure the carotid chemoreflex becomes chronically sensitised as a compensatory mechanism, and is associated with a worse prognosis^20^, exertional dyspnoea, dysfunctional or inefficient breathing, and poor exercise tolerance^21^, similar to symptoms in patients with long COVID who do not have heart failure.

SARS-CoV-2 infects host cells via binding of its receptor-binding domain to the membrane bound angiotensin-converting enzyme 2 (ACE2) ^22^. ACE2 is abundant in the carotid bodies^23^. A role of the carotid chemoreflex in the acute phase of the infection is supported by reports of silent hypoxia^23,24^, SARS-CoV-2 invasion of glomus cells (main oxygen sensing cells) and microembolism within the small arteries suppling blood to the carotid body^25,26^. The carotid bodies express their own renin-angiotensin system^27,28^, where normal functioning is dependent on the balance of ACE1 and ACE2^27^. Disruption of this local system causes increased carotid chemoreflex activity ^28,29^. Thus, disturbances in the carotid body following SARS-CoV-2 infection; by viral invasion, blood flow disruption and local immune responses could cause chemoreceptive dysfunction, by increasing local ACE1/ACE2 imbalance (in favour of higher ACE1 expression versus ACE2) and angiotensin II receptor stimulation^25,30^.

We propose that carotid body dysfunction occurs in long COVID, which contributes to dysregulation of ventilation and cardiovascular control, especially during exercise. Therefore, we conducted a case-control study to determine whether carotid chemoreflex sensitivity is elevated in non-hospitalised patients with long COVID and whether this could help to explain impairments in exercise tolerance and dysregulated breathing reported during exercise in non-hospitalised patients with ongoing symptoms. We hypothesised that long COVID patients would exhibit increased hypoxic ventilatory responses at rest and poorer ventilatory efficiency during exercise compared to controls.

## Results

### Participants

Supplementary Figure 1 shows recruitment, screening, cases excluded and the final sample size. Sixty-four individuals contacted the group about the study, and all were sent a participant information sheet. Of these, 32 replied and completed phone screening. Six were excluded due to screen failure (met exclusion criteria). Twenty-six participants were recruited (long COVID n=16 and controls n=10). Two participants (1 long COVID and 1 control) did not complete the study since they could not tolerate the mask for spirometry, 1 long COVID participant was excluded during the study visit due to identification of cardiac disease (valve disease or non-benign arrythmia) and 1 participant was excluded between visits due to diagnosis of gout (control). The final sample size was 14 participants with long COVID and 8 controls (recovered from initial viral infection within 4 weeks without ongoing symptoms). Data sets from six healthy participants from a previous study (NHS REC numbers: 17/SW/0171 and 18/SW/0241), completed before the SARS-CoV-2 pandemic using the same methods, equipment, and location (clinical room) were added to the control group. These participants were identified based on age, sex, and body mass index (BMI) so that they matched the participants in the long COVID study.

Participant demographics are shown in Table 1. Age, BMI, height, and body mass were similar between the control and long COVID groups (P>0.05). Clinic systolic blood pressure (SBP), diastolic BP (DBP) and heart rate (HR), were not different between groups (P>0.05).

**Table 1.** Demographic, spirometry, and SARS-CoV-2 infection data.

| **Demographics** | **Controls (n=14)** | **Long COVID (n=14)** | **P-value** |
| --- | --- | --- | --- |
| **Sex (M/F)** | 3/11 | 2/12 | - |
| **Age (years)** | 35 ± 13 | 42 ± 12 | 0.1195 |
| **BMI (kg/m<sup>2</sup>)** | 24.5 ± 4.9 | 26.9 ± 5.3 | 0.2140 |
| **Height (cm)** | 166 ± 5 | 167 ± 7 | 0.5543 |
| **Body mass (kg)** | 67.2 ± 12.9 | 75.1 ± 16.3 | 0.1573 |
| **Received all recommended COVID-19 vaccinations at time of study (%)** | 100% | 100% | - |
| **Time since initial infection (days) [median (IQR)]** | 473 (265-543) | 534 (400-709) | 0.1006* |
| **Clinic BP** |  |  |  |
| **SBP (mmHg)** | 121 ± 9 | 131 ± 17 | 0.0732 |
| **DBP (mmHg)** | 75 ± 7 | 83 ± 13 | 0.0594 |
| **HR (beats/min)** | 69 ± 12 | 75 ± 12 | 0.1895 |
| **Resting lung function** |  |  |  |
| **n** | 11 | 14 |  |
| **FEV<sub>1</sub> (z-scores)** | -0.32 ± 0.92 | -0.90 ± 1.23 | 0.2109 |
| **FVC (z-scores)** | -0.56 ± 1.01 | -1.03 ± 0.86 | 0.2279 |
| **FEV<sub>1</sub>/FVC (z-scores)** | 0.60 ± 0.95 | 0.21 ± 1.13 | 0.3727 |
| **FEF<sub>75%</sub> (z-scores)** | 0.61 ± 1.01 | 0.43 ± 0.77 | 0.6191 |

All participants who had COVID-19 were first infected from April 2020 to December 2021, when the dominant variants of SARS-CoV-2 were the original virus, Alpha, Delta, and Omicron. The average number of days since first infection to the study visit, in the long COVID and control groups, were similar (Table 1, P=0.1006). As per the exclusion criteria, none of the participants were hospitalised (admitted) for COVID-19 and had a previous positive PCR test at the time of the initial infection or were seropositive for SARS-CoV-2 antibodies (see inclusion/exclusion criteria in methods). The initial infection caused mild to moderate symptoms in all participants. Table 2 outlines the most common ongoing symptoms previously reported^9,31,32^ and the percentage of the long COVID participants in this study reporting these symptoms. Long COVID participants reported at least 3 symptoms, with the most common symptoms being dyspnoea at rest and on exertion, extreme fatigue, “brain fog” and chest pain.

**Table 2.** Common ongoing symptoms reported in post COVID-19 syndrome and the % of participants in the present study, self-reporting these symptoms.

| Symptoms | Long COVID (n=14) |
| --- | --- |
| At least 3 symptoms | 100% |
| Dyspnoea on exertion | 92% |
| Dyspnoea at rest | 86% |
| Brain fog | 86% |
| Fatigue | 79% |
| Chest pain | 50% |
| Palpitations or tachycardia | 36% |
| Dizziness | 36% |
| Post-exertional malaise | 36% |
| Muscle or joint pain | 21% |
| Oedema in extremities | 14% |
| Nausea | 14% |
| Chronic dry cough | 14% |
| Impaired taste or smell | 7% |
| Alopecia | 7% |
| Chronic fever | 0% |
| Diarrhoea | 0% |

Ten out of the 14 long COVID participants had been seen in a cardiology outpatient long COVID clinic at a UK based University Hospital NHS Trust, where structural cardiac disease (e.g., myocarditis, heart failure, ischaemia) had been excluded. All the participants reporting chest pain were reviewed in a cardiology clinic, which was diagnosed as non-cardiac in origin. None of the long COVID participants (or controls) had been diagnosed with pre-existing cardiovascular or pulmonary diseases. Four of the long COVID participants had been prescribed ivabradine in the cardiology outpatient long COVID clinic. These participants stopped taking ivabradine 48 hours before their study visits (plasma half-life: 2 hours and effective half-life: 11 hours).

Severe asthma was an exclusion criterion, however 2 controls and 6 long COVID participants had been previously diagnosed with mild asthma (before COVID). All participants had a normal resting 12-lead ECG, which was completed as part of the study screening process. Twelve of the 14 long COVID participants self-reported having a chest x-ray, all of which were normal. Finally, Supplementary Table 1 provides information of drugs prescribed in both groups.

### Resting spirometry

Spirometry data are summarized in Table 1 and are presented as z-scores with a normal z-score being between ± 1.64^33^. There were no differences between the two groups for FEV_1_ (forced expiratory volume in 1 second), FVC (forced vital capacity), FEV_1_/FVC and FEF_75%_ (forced expiratory flow after 75% of the FVC has been exhaled; P>0.05). No participants had an FEF_75%_ smaller than −1.64 z-score, therefore excluding small airway disease. One control subject and 3 long COVID participants had a reduced FEV_1_ and FVC with a normal FEV_1_/FVC ratio, suggestive of a restrictive ventilatory defect. One long COVID participant had a reduced FEV_1_ and FVC, with an FEV_1_/FVC ratio consistent with an obstructive ventilatory defect (z-score = −3.25) indicating severe airflow obstruction (z-score = −3.85)^33^.

### Sit to standing blood pressure test

To check for orthostatic intolerance in both groups, a sit-to-standing test (like that used in the HYVET study^34,35^) was completed (3 mins of standing). Sit-to-stand was completed in all long COVID participants and 8 of the controls. BP was measured at seated rest, immediately after standing, 1 min, 2 mins and 3 mins of standing.

According to HYVET study guidelines, a fall in SBP>15 and DBP>7 mmHg was used as a diagnosis of orthostatic intolerance. During standing, the mean SBP and DBP did not fall by more than 15 and 7 mmHg in either group at any time point ^34^ (Supplementary Figure 2; Supplementary Table 2 for mean change data). Two control participants had a fall in SBP >15 or DBP >7 mmHg, and 1 participant from the long COVID group met this criterion (supplementary figure 2). Mean data at each timepoint were analysed using a mixed model ANOVA. Interestingly, we found that the increase in HR was larger in the control group versus the long COVID group (main Time*Group effect; P=0.0258, see supplementary results for ANOVA details and figure 2).

### Resting cardiopulmonary data

The resting HR, BP, minute ventilation, tidal volume and breathing frequency were similar between groups (Table 3, P>0.05). Resting partial pressure of the end tidal CO_2_ (P_ET_CO_2_) was lower in the long COVID group (range: 22 to 34 mmHg versus controls (range: 30-36 mmHg, Table 3, P=0.0378). The minute ventilation was similar between groups despite a lower P_ET_CO_2_ in the long COVID group. This was coupled with a higher ventilation for the volume of CO_2_ expired (V_E_/VCO_2_ ratio, P=0.0246); suggesting that there is some hyperventilation occurring at rest in the long COVID group. There were no differences in the expiratory and inspiratory times, or the ratio of inspiration to expiratory times between groups (P>0.05; Table 3 for exact P-values). Interestingly, the tidal volume/inspiratory time ratio (V*_T_*/Ti; an index of inspiratory flow^36^) was 16% higher in the long COVID group versus the controls (P=0.0483) indicative of increased inspiratory drive.

**Table 3.**
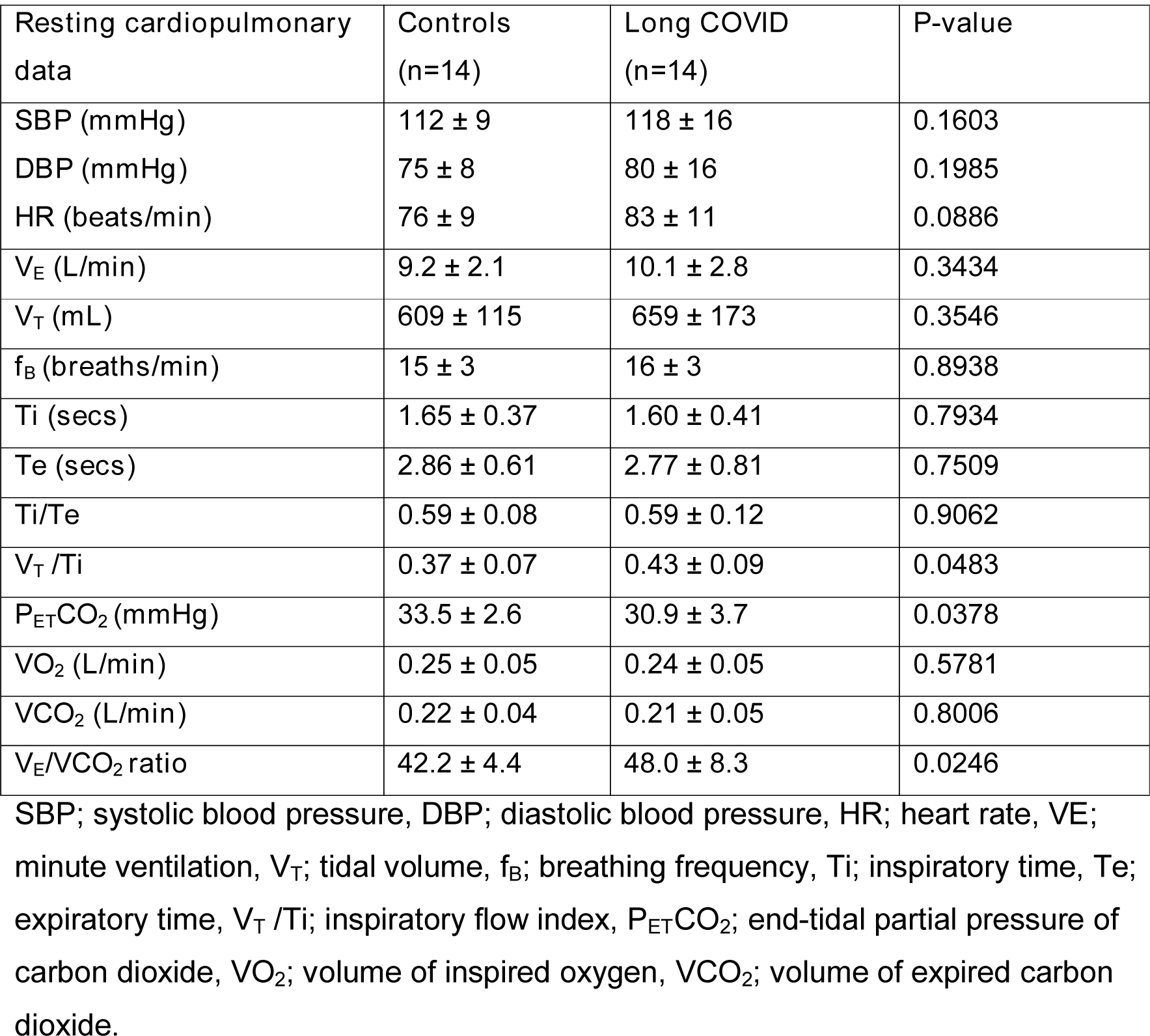
Resting cardiopulmonary variables in controls versus long COVID.

| Resting cardiopulmonary data | Controls (n=14) | Long COVID (n=14) | P-value |
| --- | --- | --- | --- |
| SBP (mmHg) | 112 ± 9 | 118 ± 16 | 0.1603 |
| DBP (mmHg) | 75 ± 8 | 80 ± 16 | 0.1985 |
| HR (beats/min) | 76 ± 9 | 83 ± 11 | 0.0886 |
| $V_E$ (L/min) | 9.2 ± 2.1 | 10.1 ± 2.8 | 0.3434 |
| $V_T$ (mL) | 609 ± 115 | 659 ± 173 | 0.3546 |
| $f_B$ (breaths/min) | 15 ± 3 | 16 ± 3 | 0.8938 |
| $Ti$ (secs) | 1.65 ± 0.37 | 1.60 ± 0.41 | 0.7934 |
| $Te$ (secs) | 2.86 ± 0.61 | 2.77 ± 0.81 | 0.7509 |
| $Ti/Te$ | 0.59 ± 0.08 | 0.59 ± 0.12 | 0.9062 |
| $V_T/Ti$ | 0.37 ± 0.07 | 0.43 ± 0.09 | **0.0483** |
| $P_{ET}CO_2$ (mmHg) | 33.5 ± 2.6 | 30.9 ± 3.7 | **0.0378** |
| $VO_2$ (L/min) | 0.25 ± 0.05 | 0.24 ± 0.05 | 0.5781 |
| $VCO_2$ (L/min) | 0.22 ± 0.04 | 0.21 ± 0.05 | 0.8006 |
| $V_E/VCO_2$ ratio | 42.2 ± 4.4 | 48.0 ± 8.3 | **0.0246** |

SBP; systolic blood pressure, DBP; diastolic blood pressure, HR; heart rate, $V_E$ ; minute ventilation, $V_T$ ; tidal volume, $f_B$ ; breathing frequency, $Ti$ ; inspiratory time, $Te$ ; expiratory time, $V_T/Ti$ ; inspiratory flow index, $P_{ET}CO_2$ ; end-tidal partial pressure of carbon dioxide, $VO_2$ ; volume of inspired oxygen, $VCO_2$ ; volume of expired carbon dioxide.

### Cardiopulmonary exercise testing

Cardiopulmonary exercise testing (CPET) was completed on a cycle ergometer to peak oxygen consumption (VO_2_ peak) to assess exercise tolerance and identify any breathing or cardiovascular abnormalities that might not be evident at rest. We also wanted to understand whether any dysfunctional breathing (defined here as inappropriate hyperventilation or poor breathing efficiency characterised by elevated minute ventilation/volume of CO_2_ expired (V_E_/VCO_2_) slopes^17^) during exercise could be linked to carotid chemoreflex hyperactivity in the long COVID group versus control.

To assess whether maximal effort was achieved and as an objective assessment of the quality of the test, a respiratory exchange ratio>1.15, maximum predicted HR>85%, rating of perceived exertion (RPE; 6-20 Borg Scale) of 17-20 and a plateau in VO_2_ were used. Overall, 86% of the control group and 86% of the long COVID group achieved these criteria. Thus, differences between groups are likely not due to differences in effort or quality of the CPET. In fact, the control group reported a lower RPE at peak exercise (17; 17-18 (median, IQR)) versus the long COVID group (19; 18-19, P=0.0081; Mann Whitney U). Table 4 shows the mean ± SD (range) for all CPET variables. None of the participants desaturated during the test (defined as SpO_2_%<95%, where peak SpO_2_% was similar between groups), and there were no exercise induced cardiac ischaemic changes observed on the 12-lead ECG.

**Table 4.** Cardiopulmonary exercise testing data in the control and long COVID groups.

| CPET data | Controls (n=14) | Long COVID (n=14) | P-value |
| --- | --- | --- | --- |
| HR at peak (beats/min) | 170 ± 17 | 158 ± 18 | 0.0619 |
| (% predicted) | 97 ± 6 | 92 ± 9 | 0.1326 |
| VO<sub>2</sub> at peak (L/min) | 1.78±0.57 | 1.40±0.34 | **0.0378** |
| (mL/kg/min) | 26.7±7.4 | 18.6±4.7 | **0.0015** |
| (% predicted) | 91±18 | 76±18 | **0.0340** |
| HR/%VO<sub>2</sub> slope | 1.00 ± 0.15 | 0.77 ± 0.26 | **0.0114** |
| RER at peak | 1.28 ± 0.11 | 1.27 ± 0.09 | 0.7483 |
| RPE at peak (median, IQR) | 17 (16-18) | 19 (18-19) | **0.0081\*** |
| Dyspnoea at peak (median, IQR) | 5 (4-8) | 8 (5-9) | 0.0515* |
| SpO<sub>2</sub> (%) at peak | 99 ± 1 | 99 ± 1 | 0.8966 |
| VCO<sub>2</sub> at peak (L/min) | 2.29 ± 0.83 | 1.76 ± 0.52 | **0.0477** |
| P<sub>ET</sub>CO<sub>2</sub> at peak | 39.6±4.7 | 33.6±3.8 | **0.0007** |
| O<sub>2</sub> pulse peak (mL/beat) | 10.6 ± 2.8 | 8.9 ± 2.0 | 0.0650 |
| Peak SBP (mmHg) | 177 ± 21 | 171 ± 26 | 0.5115 |
| Peak DBP (mmHg) | 88 ± 12 | 90 ± 15 | 0.6286 |
| V<sub>E</sub> at peak (L/min) | 71.5± 29.2 | 66.4±22.4 | 0.6012 |
| Breathing reserve | 43 ± 18 | 42 ± 24 | 0.8708 |
| f<sub>B</sub> at peak (breaths/min) | 36 ± 10 | 37 ± 9 | 0.8705 |
| V<sub>T</sub> at peak (L) | 1.98 ± 0.47 | 1.83 ± 0.40 | 0.3440 |
| V<sub>E</sub>/VCO<sub>2</sub> ratio at peak | 31.4 ± 4.9 | 38.7 ± 5.4 | **0.0005** |
| V<sub>E</sub>/VO<sub>2</sub> ratio at peak | 39.8 ± 7.6 | 48.4 ± 8.6 | **0.0095** |
| V<sub>E</sub>/VCO<sub>2</sub> slope | 27.7 ± 4.8 | 37.8 ± 4.4 | **0.0003** |
| Nadir V<sub>E</sub>/VCO<sub>2</sub> | 25.0 ± 2.4 | 30.1±5.0 | **0.0020** |
| VO<sub>2</sub> at AT (mL/kg/min) | 18.7 ± 5.6 | 12.6 ± 4.3 | **0.0025** |
| (% of VO<sub>2</sub> peak) | 68 ± 10 | 66 ± 13 | 0.6490 |
| P<sub>ET</sub>CO<sub>2</sub> at AT (mmHg) | 44 ± 4 | 38 ± 6 | **0.0041** |
| V<sub>E</sub>/VCO<sub>2</sub> at AT | 25.5 ± 2.4 | 30.4 ± 5.4 | **0.0042** |
| HR at AT (beats/min) | 132 ± 13 | 121 ± 15 | **0.0453** |
| (% of peak HR) | 76 ± 5 | 77 ± 8 | 0.6181 |
| $\Delta$ from rest (beats/min) | $61 \pm 18$ | $38 \pm 16$ | **0.0009** |
| VO<sub>2</sub>/WR slope | $8.2 \pm 1.2$ | $8.2 \pm 1.5$ | 0.9283 |
| Heart rate recovery 1 min<br>(beats/min) | $19 \pm 7$ | $17 \pm 5$ | 0.4635 |

The VO_2_ peak measured in L/min, mL/kg/min and as a % of peak predicted were lower in the long COVID group versus controls (Table 4; P<0.05). Absolute VO_2_ at anaerobic threshold (AT) was also lower (P=0.0025), but the % of peak VO_2_ at which the AT occurred was similar between groups (P=0.6490), indicating normal metabolic function. Along these lines there were no other indications of defects in muscle metabolic pathways indicating no muscle metabolic limitations to VO_2_ peak in either group (i.e. normal peak respiratory exchange ratio (RER) and a normal VO_2_/work rate relationship^37,38^ (Table 4). Maximum HR was lower in the long COVID group versus controls, but there was no difference between groups when HR was measured as the % of predicted maximum (P=0.1326). The slope of the HR plotted against the %VO_2_ peak was lower in the long COVID group (Table 4; P=0.0114) versus controls indicating a blunted HR response to exercise and potentially some degree of chronotropic incompetence. Heart rate at AT was also lower in the long COVID group versus controls (P=0.0453). Finally, the peak oxygen pulse (VO_2_/HR) was lower in the long COVID group versus controls, indicating a cardiovascular limitation to exercise. Despite this, the HR recovery at 1 min was similar between groups (P=0.4653); indicating that parasympathetic engagement post-exercise in the long COVID group was similar to control.

The VCO_2_ at peak was lower in the long COVID group (Table 4; P=0.0477), whereas minute ventilation, tidal volume and breathing frequency at peak were similar between groups (Table 4, P>0.05). Supplementary figure 3 shows the minute ventilation versus the VCO_2_ and VO_2_ at rest, AT, and peak exercise in both groups. The mean breathing reserve (max minute ventilation/predicted maximal voluntary ventilation (calculated as FEV1 * 40) was similar between groups (P=0.8708). Thus it is likely that the lower peak VO_2_ in the long COVID group is not a result of pulmonary mechanical limitations^39,40^.

The V_E_/VCO_2_ *ratio* at peak (ANOVA; P=0.0051) and at AT (P=0.0477) was higher in the long COVID group versus control (see Supplementary figure 3 for ANOVA details and V_E_/VCO_2_ ratio plotted against the VO_2_ at rest, AT, and peak exercise). The V_E_/VCO_2_ slope (Table 4, P=0.0008) and the V_E_/VCO_2_ nadir (P=0.0020) during exercise were also higher in the long COVID group versus controls indicating lower breathing efficiency (a higher minute ventilation to remove a given volume of CO_2_) indicating hyperventilation at any point during exercise (Figure 1 for example slopes (raw data) in a control and long COVID participant). Thirteen percent of the control group had a V_E_/VCO_2_ slope higher than the normal range (20-30^41,42^) versus 88% in the long COVID group. Finally, the P_ET_CO_2_ was lower at the AT and at peak exercise in the long COVID group versus the control group (time*group effect; P=0.0119, mixed model ANOVA), but the magnitude of rise of the P_ET_CO_2_ from rest to AT and from rest to peak exercise was the same in both groups (P=0.0904, supplementary figure 4, mixed model ANOVA).

**Figure 1.**
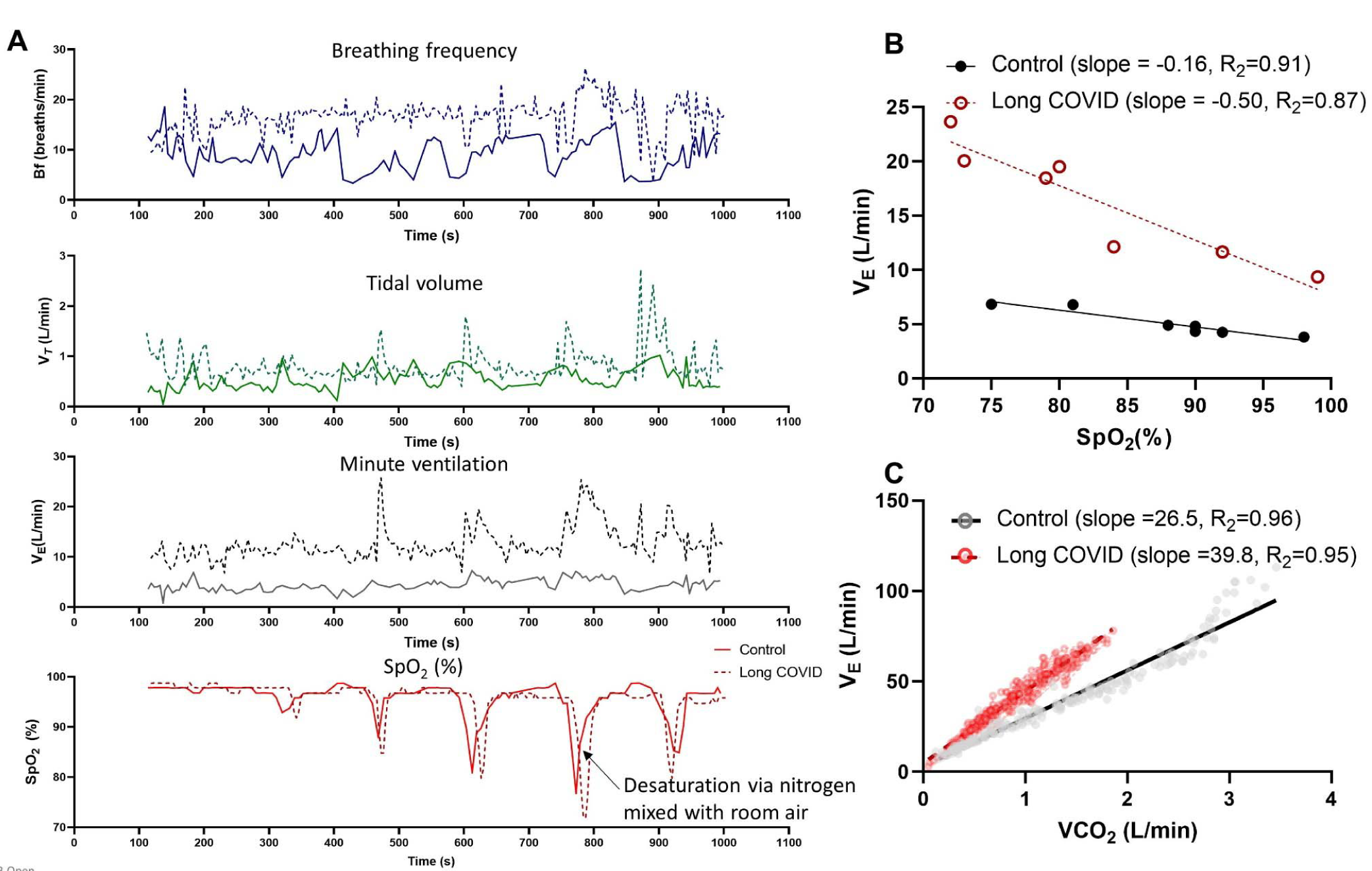
A) Example carotid chemoreflex test in a control (solid line) and long COVID (dotted line) participants. Data shows the raw SpO_2_ response to nitrogen exposure (bottom panel) and the resultant tidal volume, breathing frequency, and minute ventilation responses. The minute ventilation (average of the largest 2 consecutive breaths) is plotted against the nadir SpO_2_ for each nitrogen exposure^21^ (panel B). The resultant slope of the linear regression is the hypoxic ventilatory response. Panel C shows the minute ventilation (V_E_) plotted against the volume of expired carbon dioxide (VCO_2_) for each breath during exercise in the same 2 participants. The slope of the regression is the V_E_/VCO_2_ slope and is used as a measure of breathing efficiency.

### Hypoxic ventilatory response

Figure 1A shows examples carotid chemoreflex tests in a control and a long COVID participants and the resultant hypoxic ventilatory responses (calculated from the minute ventilation) in these participants (Figure 1B). The hypoxic ventilatory response (minute ventilation response to reductions in SpO_2_%) was elevated in the long COVID (−0.44 ± 0.23 l/min/ SpO_2_%, R^2^=0.77±0.20) group compared to controls (−0.17 ± 0.13 l/min/SpO2%, R^2^=0.54±0.38, Figure 2A, P=0.0007); thus, for a given decrease in SpO_2_, the participants with long COVID had a greater increase in minute ventilation. This was driven by a greater increase in tidal volume (Figure 2B) in the participants with long COVID rather than a greater increase in breathing frequency compared to the controls (Figure 2C). Taken together this indicates that the participants with long COVD have an elevated carotid chemoreflex sensitivity to hypoxia.

**Figure 2.**
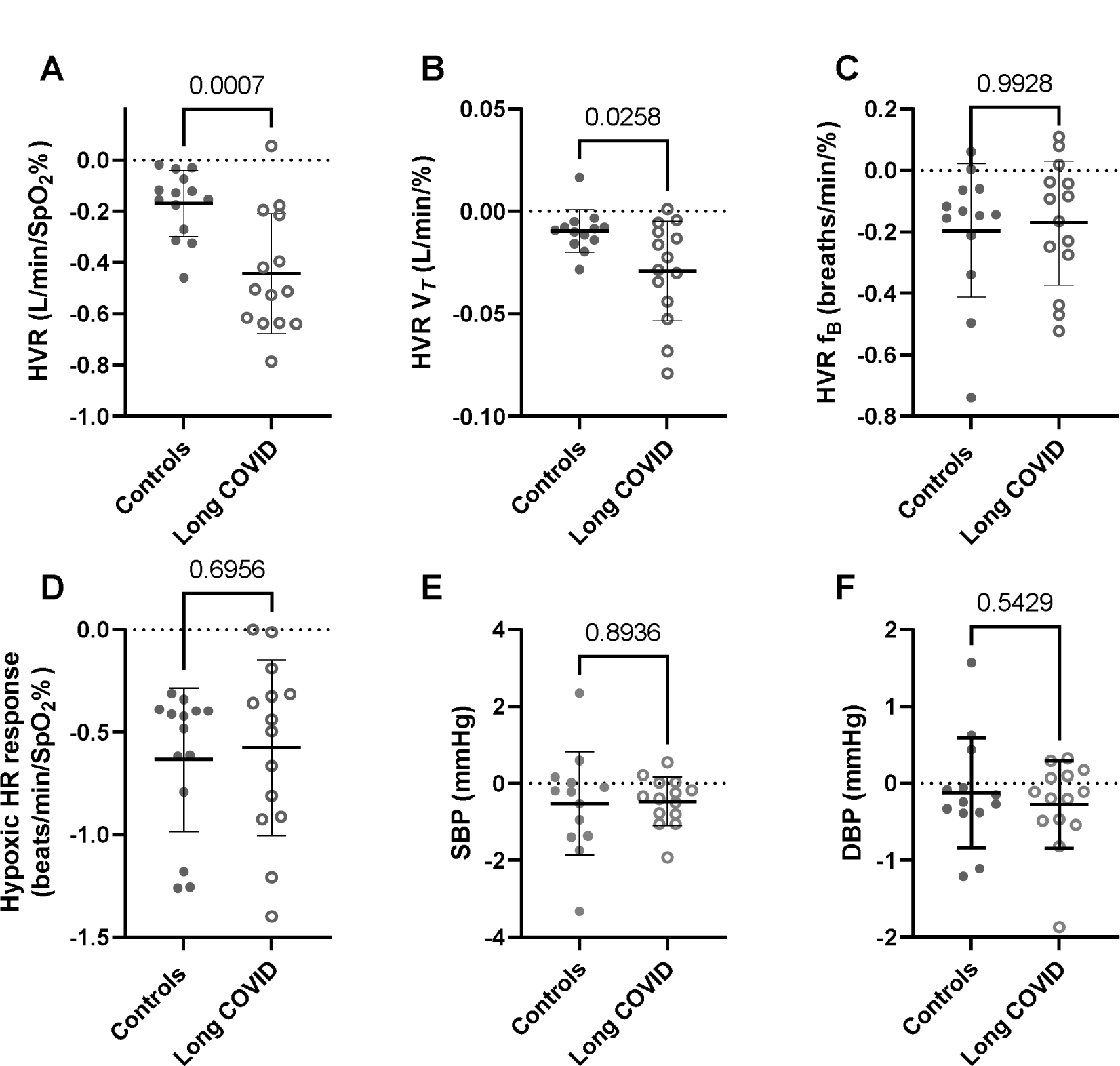
The hypoxic ventilatory response based on A) minute ventilation, B) tidal volume (V*_T_*) and C) breathing frequency (f_B_). There was a higher hypoxic ventilatory response based on minute ventilation and tidal volume in the participants with long COVID (n=14) versus control (n=14). Panels D, E and F show the heart rate, systolic and diastolic blood pressure hypoxic responses. Data are the slope of the linear regression where SpO_2_% was plotted against the heart rate, and BP during each hypoxic exposure. There was no difference in the HR or BP response to hypoxia between groups.

There was no difference in the HR response to hypoxia (Figure 2D) between groups (control; −0.66 ± 0.35 beats/min/ SpO_2_% versus long COVID; −0.58 ± 0.60 beats/min/SpO2%, P=0.5978). Additionally, there was no difference in the SBP or DBP response to hypoxia between groups (Figure 2E and 2F). The BP response to hypoxia was variable between individuals with some participants showing a depressor response to hypoxia and others showing a pressor (most common, where the slope of the regression is negative in figure 2E and 2F) or biphasic response to hypoxia.

The hypoxic ventilatory response was correlated with the V_E_/VCO_2_ slope during exercise (r=0.54, P=0.0037); indicating that the high carotid chemoreflex sensitivity in the long COVID participants may partially explain the higher V_E_/VCO_2_ slope (poorer breathing efficiency) during exercise. Therefore, the carotid chemoreflex may play an important role in driving hyperventilation or poor breathing efficiency during exercise. Targeting the carotid body or the carotid chemoreflex could be helpful in improving ongoing symptoms in participants with long COVID and improving exercise tolerance.

## Discussion

The pathophysiological mechanisms driving ongoing symptoms in patients with long COVID, after an initial mild infection, are unclear. Here we show for the first time that carotid chemoreflex sensitivity is amplified in non-hospitalised patients with long COVID (versus a control group) and that this was correlated with hyperventilation and poor breathing efficiency during exercise. Elevated carotid chemoreceptor activity could explain several of the ongoing symptoms experienced by patients living with long COVID.

### The hyperventilatory state of long COVID

In this population of participants with long COVID, we show that despite similar lung function (resting spirometry and ventilatory reserve during exercise) to the control group, these participants hyperventilate at rest and during exercise. At rest, this is evidenced by a similar minute ventilation compared to control despite a lower resting P_ET_CO_2_. Resting levels of arterial CO_2_ provides the key stimulus for respiratory drive, mainly via the central chemoreceptors.^43^ Reductions in P_ET_CO_2_ indicate decreases in PaCO_2_ ^44,45^ which is a stimulus to lower ventilation; however, this has not occurred in the long COVID participants since their level of ventilation was the same as the controls suggesting a resetting of chemoreceptor set-point for breathing. If resting P_ET_CO_2_ levels were restored to normal control levels, it is possible that minute ventilation would be greater in the long COVID group. Additionally, the V_E_/VCO_2_ ratio at rest is higher in the long COVID participants, showing that they are breathing more to remove the same volume of CO_2_ as controls.

Altered V_E_/VCO_2_ ratio (and P_ET_CO_2_) could result from changes to gas exchange between the alveoli and pulmonary circulation. However, we found that during exercise the magnitude of the rise in P_ET_CO_2_ was the same in the long COVID participants versus the controls. During exercise (with a normal gas exchange) P_ET_CO_2_ rises and peaks around anaerobic threshold. When gas exchange is impaired by abnormal ventilation or perfusion of the lung, the P_ET_CO_2_ will show smaller rises, no rise, or even decrease as seen in COPD, heart failure and pulmonary hypertension^46^. As such, the similar rise in P_ET_CO_2_ with exercise in long COVID patients and control participants suggests no differences in gas exchange between these groups. However, we acknowledge that this needs to be confirmed with gas diffusion studies.

We also found evidence of elevated resting V*_T_*/Ti, an index of inspiratory flow, in the long COVID group versus the control group. The V*_T_*/Ti is used as surrogate of primary neural drive for inspiration^36^ and thus could indicate a higher ‘inspiratory drive’ at rest in the long COVID participants which may contribute to their feelings of dyspnoea. High inspiratory flow can also indicate changes in airway mechanics^47^, however, this is unlikely in the long COVID group, since resting lung function (spirometry) was similar between groups. During exercise, hyperventilation or poor breathing efficiency was also evidenced by elevated V_E_/VCO_2_ slopes and an elevated V_E_/VCO_2_ ratio at anaerobic threshold and peak exercise. Since the carotid chemoreflex plays an important part in the control of breathing both at rest and exercise (including inspiratory drive^48^); it is possible that it is contributing to hyperventilation in long COVID.

### Carotid chemoreflex

The mammalian carotid chemoreflex is a protective reflex that contributes to ventilatory and cardiovascular control^49^. In addition to signalling the need for increased ventilation during hypoxia (and other stimuli^19^), the carotid body chemoreceptors contribute to resting ventilatory drive, as demonstrated by their tonic activity in humans and animal models^48,50^. It is well established that certain disease states exhibit exaggerated carotid chemoreflex sensitivity^20^ and tonicity^18^. Given that elevated carotid chemoreflex sensitivity measured by the hypoxic ventilatory response predicts symptom burden, exercise intolerance, dyspnoea, and hyperventilation in heart failure^21,51^, we aimed to assess carotid chemoreflex sensitivity in long COVID. The hypoxic ventilatory response was 145% higher in the long COVID group versus age-, BMI- and sex-matched controls indicating amplified carotid chemoreceptor sensitivity in the long COVID group. The higher hypoxic ventilatory response was observed despite a lower resting P_ET_CO_2_ in the long COVID group even though hypocapnia normally depresses carotid chemoreflex sensitivity to hypoxia^52^. The hypoxic ventilatory response was in fact similar to that measured in a group of patients with heart failure reduced ejection fraction in our lab (−0.44 ±0.23 L/min/SpO2% versus −0.48± 0.30 L/min/SpO2%; mean ± SD) using the same equipment before March 2020 (see supplementary Figure 5). The elevated hypoxic ventilatory response in the long COVID participants was driven by amplified responses in tidal volume rather than breathing frequency, which supports previous reports in humans where the increase in minute ventilation during mild hypoxia is driven mainly by increases in tidal volume^53^.

The hypoxic ventilatory response was correlated with the V_E_/VCO_2_ slope, so that a person with high chemoreflex sensitivity had a higher V_E_/VCO_2_ slope (or vice versa). This suggests that elevated carotid chemoreflex sensitivity may partially explain reduced breathing efficiency and hyperventilation during exercise in participants with long COVID^14,15,17^. In fact, breathing efficiency was poor in some of the participants in this study; 64% of the long COVID group had a V_E_/VCO_2_ slope >34 (versus only 7% (one person) in the control group), which is a powerful prognostic indicator of future outcomes for people with heart failure^54^. It is possible that in long COVID the carotid chemoreflex has a similar function as that described heart failure, partially explaining the dysregulated breathing and feelings of breathlessness in patients with ongoing symptoms after their initial mild infection. Elevated V_E_/VCO_2_ slopes are also caused by increased dead space ventilation due to 1) inadequate aeration of the alveoli and 2) poor perfusion of the aerated lung spaces, affecting gas exchange. However, in this study there was no evidence of mechanical lung or small airway issues (due to a similar prevalence of normal breathing reserve in the two groups and similar resting spirometry between the groups). No obvious evidence of gas exchange issues were observed either, because the P_ET_CO_2_ during exercise (to AT) increased by a similar magnitude in the participants with long COVID versus the controls.

Finally, the resting P_ET_CO_2_ was <30 mmHg in 4 participants with long COVID. Low P_ET_CO_2_ causes similar symptoms as those often experienced by patients with long COVID including feelings of brain fog (due to poor cerebral perfusion) and paraesthesia ^55^. The level of P_ET_CO_2_ however did not reach apnoeic threshold levels ^56^ in any patients because there were no apnoeas at rest and no evidence of periodic breathing patterns^57^. Low resting P_ET_CO_2_ and hyperventilation is a hallmark of hyperventilatory syndrome which has been cited as a cause of ongoing in symptoms in some cohorts of patients with long COVID^58^. It is possible that problems with breathing at rest and during exercise, leading to lower P_ET_CO_2_ levels and manifesting as hyperventilation syndrome could be partially driven by the carotid chemoreflex in patients with long COVID.

### Mechanisms

Possible mechanisms of increased carotid chemoreceptor sensitivity after SARS-CoV-2 infection include local changes within the parenchyma of the carotid body and/or dysfunction occurring in medullary regions that process afferent sensory information and control efferent ventilatory responses.

In the acute phase of the infection, local viral and immune cell invasion of the carotid body^25^ could disrupt normal functioning, potentially via immune cells destroying infected glomus cells explaining silent hypoxia in some patients^23^. In the long-term these cells are likely replaced but there is the possibility of long-term local inflammation and disruption of ACE1 and ACE2 balance leading to elevated carotid chemoreceptor drive. Additionally, since the carotid bodies are highly sensitive to disturbances in perfusion, any blood flow disruption caused by microthrombi and endothelial dysfunction^25^ could elevate carotid body activity^59^. It is also possible that dysfunction of the petrosal ganglion, which carries afferent sensory input into the brainstem, causes hyper-reactivity in the carotid chemoreflex. Of note, the petrosal ganglion participates in mediating taste signals in the brain. However, only 7% of our long COVID participants reported ongoing impaired sense of taste and smell ^11^.

Additionally, inflammation or neuronal damage in the medullary regions mediating afferent signals from the carotid body could contribute to increased chemoreceptor drive. Along these lines there is evidence from animal models that the original SARS-CoV infiltrates medullary brain regions^60^, but there is no strong evidence of this occurring with existing variants of SARS-CoV-2.

Finally, some evidence suggests the SARS-CoV-2 may affect mitochondrial function^61^. The glomus cells in the carotid body have unique mitochondria, which are important for O_2_ sensing^62^. Any mitochondrial dysfunction in glomus cells could lead to augmented chemoreflex activity^62^. Since mitochondrial dysfunction could occur in any organ, the exercise intolerance and hyperventilation observed in long COVID could also be driven, for example, by the metaboreceptors in skeletal muscle due to poor mitochondrial function and this needs to be examined.

### Limitations

Firstly, the control group are a combination of prospectively recruited participants who recovered from a SARS-CoV-2 infection in <4 weeks, and participants who were retrospectively identified from our previous studies (all measures were taken using the same equipment, procedures, and site). Thus 6 of the controls did not have a previous SARS-CoV-2 infection. We cannot rule out that SARS-CoV-2 infection has long lasting effects of the cardiopulmonary system even if individuals recover with no ongoing symptoms. Secondly, we measured the carotid chemoreflex sensitivity using poikilocapnic hypoxia, thus P_ET_CO_2_ decreased after each hypoxic exposure (due to ventilatory adjustments). However, it is unlikely that this affected the ventilatory response to hypoxia because 1) the P_ET_CO_2_ decreased only after the breaths that were used as measurements for ventilation and thus could not affect the data and 2) we waited for the P_ET_CO_2_ to return to baseline after each hypoxic exposure. Thirdly, this was a single site study with a small sample size and thus needs to be evaluated in a larger population of patients with long COVID. Finally, although we see evidence of elevated carotid chemoreceptor sensitivity in patients with long COVID, we need to evaluate whether dampening down the hyperreflexia helps to improve symptoms.

### Implications

Here we show for the first time that patients with long COVID have elevated carotid chemoreceptor sensitivity and that this is correlated with poor breathing efficiency or hyperventilation during exercise. Interventions that temper carotid body excitability could be explored as a treatment option for long COVID. Previously our group had shown that P2X3 receptors in the carotid body can be targeted to reduce carotid chemoreflex hyperreflexia in an animal model of hypertension^18^ and heart failure^63^, and could be a viable target in humans with long COVID. Gefapixant, an oral P2X3 receptor antagonist, has recently demonstrated efficacy and an acceptable safety profile in chronic cough in phase 3 clinical trials^64^. P2X3 receptors could therefore be a viable target in humans with long COVID.

## Methods

### Design

This was a single-site case-control study.

### Participants

Ethical approval for the study was granted by South Central Hampshire NHS Research Ethics Committee (21/SC/0260) and the Health Research Authority. Participants gave their written informed consent. All participants were asked to abstain from intense exercise and alcohol consumption 24 hours before the study. All experimental protocols conformed to the Declaration of Helsinki. Inclusion criteria for all participants were; aged 18-80 years and a positive SARS-CoV-2 antibody test before vaccination, or a positive COVID-19 PCR antigen swab test. Long COVID participants had received a diagnosis of long COVID, where symptoms developed during or after an infection consistent with COVID-19 and continued for more than 12 weeks (and could not be explained by an alternative diagnosis) as per NHS (UK) National Institute for Health and Care Excellence guidelines^65^.

Participants without long COVID had symptoms lasting less than 4 weeks after their initial infection. See the online-only Supplement for exclusion criteria.

### Experimental protocol

Participants attended the NIHR Bristol Clinical Research facility for 2 studies, completed at the same time of day, and the laboratory conditions were at a set temperature (22°C). In visit one, informed consent, in-depth medical history, clinic blood pressure assessment, sit-to-stand test for orthostatic intolerance, lung function tests (spirometry), 12-lead ECG, pregnancy tests and a symptom limited incremental cardiopulmonary exercise test were completed. The second visit involved resting ventilation and cardiovascular measurements followed by carotid chemoreflex assessment via the hypoxic ventilatory response.

### Procedures

*Clinic blood pressure:* Participants rested in a chair for 10 minutes prior to clinic BP being assessed (Omron, 705IT, Omron Healthcare, Kyoto, Japan). Clinic BP was assessed in-line with European Hypertension Society Guidelines^66^

*Orthostatic intolerance* was assessed using a sit-to-stand test using the HYVET protocol^34^ where BP was measured when sitting, immediately upon standing, and after 1, 2 and 3 minutes of standing.

*Resting spirometry:* Resting spirometry was used to assess lung function, to ensure no mechanical lung function abnormalities were present. Spirometry (Ergostick, CPET system, LoveMedical, UK) was completed in line with the joint American Thoracic and European Respiratory Society guidelines ^67^. The Global Lung Function Initiative (GLI) network reference values were used to calculate the percentage of predicted values and z-scores^68^.

*12-lead ECG:* Resting 12-lead ECG was performed and checked by a Cardiologist at the Bristol Heart Institute for any ECG abnormalities, and to clear participants to exercise.

### Cardiopulmonary exercise testing

After acclimatisation to the facemask and sitting on the cycle ergometer, participants completed a 5-minute steady state resting period followed by 3 minutes of unloaded cycling. Participants then completed a continuous ramp incremental exercise test to volitional exhaustion where work rate increased by 15-30 W depending on their physical ability.

Exercise tests were completed on an electronically braked cycle ergometer (Ergoselect 5; Ergoline, Germany). Cardiorespiratory data were recorded using a metabolic measurement system (Ergostik; LoveMedical, UK) with integrated 12-lead ECG and finger pulse oximetry for heart rate and SpO_2_% monitoring. Brachial arterial blood pressure was measured via an integrated automated auscultatory blood pressure cuff (LoveMedical, UK. Ratings of perceived exertion (6-20 Borg scale) and dyspnoea scores (modified Borg scale) were obtained at rest, and every minute during exercise, and at the end of exercise. Peak cardiopulmonary data were averaged over the last 30 seconds of exercise. The anaerobic threshold was measured via the V-slope method and Dual Criterion methods as recommended by the American Thoracic Society guidelines ^69^. The minute ventilation and VCO_2_ values from initiation to peak exercise were used to measure V_E_/VCO_2_ slope via least squares linear regression^70^.

### Carotid chemoreflex assessment

Resting carotid chemoreceptor sensitivity tests were completed with participants in a semi-supine position with continuous monitoring of beat-to-beat blood pressure (Finapres), SpO_2_(ear-lobe pulse oximeter; Radical-7; Masimo Corp, USA), heart rate (lead II of 3-lead ECG, AD Instruments, Australia). Simultaneously, ventilation was measured via a facemask attached to a one-way non-rebreathing circuit (Hans Rudolph, Inc., USA). The inhalation part of the circuit delivered room air or 100% nitrogen gas (transiently) for carotid chemosensitivity testing. The exhalation arm of the circuit was connected to a gas analyser (Ad Instruments) and flow head (MLT3000L; AD Instruments) fitted with a differential pressure transducer (FE141 Spirometer; AD Instruments) for the measurement of inspired and expired fractions of O_2_ and CO_2_, tidal volume, breathing frequency and minute ventilation. All data were continuously monitored and recorded with a data acquisition system (Powerlab 16/30; AD Instruments) and stored for subsequent analyses using associated software (LabChart 8.0 Pro; AD Instruments).

The transient hypoxic ventilatory response test was used to measure the sensitivity of the carotid chemoreflex ^21,71^. After a period of quiet rest breathing room air (10 mins baseline), the researcher added extra-nitrogen to the room air being delivered to the face-mask.

Nitrogen gas administration was controlled silently using a high-pressure electric valve. The nitrogen blended into the room air was delivered for 2-8 breaths, followed by a 3-minute recovery period or until ventilation and haemodynamic variables returned to baseline levels. This was repeated 6-8 times to obtain a range of oxygen saturations (SpO_2_: ∼70–100%).

The average of the two largest consecutive breaths in the 1 minute proceeding the nitrogen exposure was used to calculate the ventilatory response to reductions in SpO2%^20^. The hypoxic ventilatory response was evaluated as the slope of the linear regression relating the minute ventilation to the nadir of SpO_2_ for each nitrogen exposure ^20,71^ (Figure 1B). The response of tidal volume and breathing frequency to reductions in SpO_2_ were also evaluated in the same way as that for minute ventilation.

The peak heart rate and blood pressure was determined following each hypoxic challenge using a 3-beat rolling average and plotted against the nadir oxygen saturation. The hypoxic heart rate (beats/min/%) and blood pressure (mmHg/%) were calculated as the slope of the simple linear regression obtained from baseline and the hypoxic challenges.

### Statistical analysis

Statistical analyses were completed in GraphPad Prism (V9.5.1). Participant demographics, resting spirometry, cardiopulmonary exercise and hypoxic ventilatory response data were analysed using an independent samples t-test or Mann Whitney U test if data were not normally distributed or were non-parametric. Where data are compared across multiple time points between the groups, a mixed model ANOVA was used with a Bonferroni correction for pairwise comparisons. Data are reported as mean ± standard deviation or median (interquartile range) unless otherwise stated. A significance level of α<0.05 was used for all analyses.

## Data Availability

All data produced in the present study are available upon reasonable request to the author

## Acknowledgements

Thank you to the participants who took part (despite their often-debilitating symptoms), the study would not have been possible without them. Thank you also to the cardiology research nurses who helped support this study.

## References

1. Venkatesan, P. NICE guideline on long COVID. Lancet Respir Med 9, 129 (2021).

2. Thompson, E.J., et al. Long COVID burden and risk factors in 10 UK longitudinal studies and electronic health records.

3. Ahmad, I., et al. High prevalence of persistent symptoms and reduced health-related quality of life 6 months after COVID-19. Front Public Health 11, 1104267 (2023).

4. Augustin, M., et al. Post-COVID syndrome in non-hospitalised patients with COVID-19: a longitudinal prospective cohort study. Lancet Reg Health Eur 6, 100122 (2021).

5. O’Mahoney, L.L., et al. The prevalence and long-term health effects of Long Covid among hospitalised and non-hospitalised populations: a systematic review and meta-analysis. eClinicalMedicine 55, 101762 (2023).

6. Huang, C., et al. 6-month consequences of COVID-19 in patients discharged from hospital: a cohort study. Lancet 397, 220–232 (2021).

7. Antonelli, M., et al. Risk factors and disease profile of post-vaccination SARS-CoV-2 infection in UK users of the COVID Symptom Study app: a prospective, community-based, nested, case-control study. Lancet Infect Dis 22, 43–55 (2022).

8. (ONS), O.f.N.S. Prevalence of ongoing symptoms following coronavirus (COVID-19) infection in the UK: 2 February 2023. (ONS website, statistical bulletin, 2023).

9. Whitaker, M., et al. Persistent COVID-19 symptoms in a community study of 606,434 people in England.

10. Ballering, A.V., van Zon, S.K.R., Olde Hartman, T.C. & Rosmalen, J.G.M. Persistence of somatic symptoms after COVID-19 in the Netherlands: an observational cohort study. Lancet 400, 452–461 (2022).

11. Alkodaymi, M.S., et al. Prevalence of post-acute COVID-19 syndrome symptoms at different follow-up periods: a systematic review and meta-analysis. Clin Microbiol Infect 28, 657–666 (2022).

12. Davis, H.E., McCorkell, L., Vogel, J.M. & Topol, E.J. Long COVID: major findings, mechanisms and recommendations. Nat Rev Microbiol 21, 133–146 (2023).

13. Davis, H.E., McCorkell, L., Vogel, J.M. & Topol, E.J. Long COVID: major findings, mechanisms and recommendations. Nature Reviews Microbiology 21, 133–146 (2023).

14. Durstenfeld, M.S., et al. Use of Cardiopulmonary Exercise Testing to Evaluate Long COVID-19 Symptoms in Adults: A Systematic Review and Meta-analysis. JAMA Netw Open 5, e2236057 (2022).

15. Singh, I., et al. Persistent Exertional Intolerance After COVID-19: Insights From Invasive Cardiopulmonary Exercise Testing. Chest 161, 54–63 (2022).

16. van Voorthuizen, E.L., van Helvoort, H.A.C., Peters, J.B., van den Heuvel, M.M. & van den Borst, B. Persistent Exertional Dyspnea and Perceived Exercise Intolerance After Mild COVID-19: A Critical Role for Breathing Dysregulation? Phys Ther 102(2022).

17. Frésard, I., et al. Dysfunctional breathing diagnosed by cardiopulmonary exercise testing in ‘long COVID’ patients with persistent dyspnoea. BMJ Open Respir Res 9(2022).

18. Pijacka, W., et al. Purinergic receptors in the carotid body as a new drug target for controlling hypertension. Nat Med 22, 1151–1159 (2016).

19. Iturriaga, R., Alcayaga, J., Chapleau, M.W. & Somers, V.K. Carotid body chemoreceptors: physiology, pathology, and implications for health and disease. Physiol Rev 101, 1177–1235 (2021).

20. Ponikowski, P., et al. Peripheral chemoreceptor hypersensitivity: an ominous sign in patients with chronic heart failure. Circulation 104, 544–549 (2001).

21. Chua, T.P., et al. Clinical characteristics of chronic heart failure patients with an augmented peripheral chemoreflex. Eur Heart J 18, 480–486 (1997).

22. Lan, J., et al. Structure of the SARS-CoV-2 spike receptor-binding domain bound to the ACE2 receptor. Nature 581, 215–220 (2020).

23. Villadiego, J., et al. Is Carotid Body Infection Responsible for Silent Hypoxemia in COVID-19 Patients? Function (Oxf) 2, zqaa032 (2021).

24. Villadiego, J., et al. Is Carotid Body Infection Responsible for Silent Hypoxemia in COVID-19 Patients? Function 2(2020).

25. Porzionato, A., et al. Case Report: The Carotid Body in COVID-19: Histopathological and Virological Analyses of an Autopsy Case Series. Front Immunol 12, 736529 (2021).

26. Lambermont, B., Davenne, E., Maclot, F. & Delvenne, P. SARS-CoV-2 in carotid body. Intensive Care Med 47, 342–343 (2021).

27. Schultz, H.D. Angiotensin and carotid body chemoreception in heart failure. Curr Opin Pharmacol 11, 144–149 (2011).

28. Allen, A.M. Angiotensin AT1 receptor-mediated excitation of rat carotid body chemoreceptor afferent activity. J Physiol 510 (Pt 3), 773–781 (1998).

29. Li, Y.L., et al. Angiotensin II enhances carotid body chemoreflex control of sympathetic outflow in chronic heart failure rabbits. Cardiovasc Res 71, 129–138 (2006).

30. Rysz, S., et al. COVID-19 pathophysiology may be driven by an imbalance in the renin-angiotensin-aldosterone system. Nat Commun 12, 2417 (2021).

31. Shah, W., Hillman, T., Playford, E.D. & Hishmeh, L. Managing the long term effects of covid-19: summary of NICE, SIGN, and RCGP rapid guideline. BMJ 372, n136 (2021).

32. Cervia, C., et al. Immunoglobulin signature predicts risk of post-acute COVID-19 syndrome. Nature Communications 13, 446 (2022).

33. Sylvester, K.P., et al. ARTP statement on pulmonary function testing 2020. BMJ Open Respir Res 7(2020).

34. Peters, R., et al. Orthostatic hypotension and symptomatic subclinical orthostatic hypotension increase risk of cognitive impairment: an integrated evidence review and analysis of a large older adult hypertensive cohort. Eur Heart J 39, 3135–3143 (2018).

35. Peters, R., et al. Orthostatic hypotension and symptomatic subclinical orthostatic hypotension increase risk of cognitive impairment: an integrated evidence review and analysis of a large older adult hypertensive cohort. European Heart Journal 39, 3135–3143 (2018).

36. Morgan, B.J., Adrian, R., Bates, M.L., Dopp, J.M. & Dempsey, J.A. Quantifying hypoxia-induced chemoreceptor sensitivity in the awake rodent. Journal of applied physiology 117, 816–824 (2014).

37. Riley, M.S., Nicholls, D.P. & Cooper, C.B. Cardiopulmonary Exercise Testing and Metabolic Myopathies. Ann Am Thorac Soc 14, S129–S139 (2017).

38. Riley, M.S., Nicholls, D.P. & Cooper, C.B. Cardiopulmonary Exercise Testing and Metabolic Myopathies. Annals of the American Thoracic Society 14, S129–S139 (2017).

39. Glaab, T. & Taube, C. Practical guide to cardiopulmonary exercise testing in adults. Respir Res 23, 9 (2022).

40. Glaab, T. & Taube, C. Practical guide to cardiopulmonary exercise testing in adults. Respiratory Research 23, 9 (2022).

41. Balady, G.J., et al. Clinician’s Guide to cardiopulmonary exercise testing in adults: a scientific statement from the American Heart Association. Circulation 122, 191–225 (2010).

42. Balady, G.J., et al. Clinician’s Guide to cardiopulmonary exercise testing in adults: a scientific statement from the American Heart Association. Circulation 122, 191–225 (2010).

43. Guyenet, P.G. & Bayliss, D.A. Central respiratory chemoreception. Handb Clin Neurol 188, 37–72 (2022).

44. Jones, N.L., Robertson, D.G. & Kane, J.W. Difference between end-tidal and arterial PCO2 in exercise. J Appl Physiol Respir Environ Exerc Physiol 47, 954–960 (1979).

45. Jones, N.L., Robertson, D.G. & Kane, J.W. Difference between end-tidal and arterial PCO2 in exercise. Journal of applied physiology 47, 954–960 (1979).

46. Hansen, J.E., Ulubay, G., Chow, B.F., Sun, X.G. & Wasserman, K. Mixed-expired and end-tidal CO2 distinguish between ventilation and perfusion defects during exercise testing in patients with lung and heart diseases. Chest 132, 977–983 (2007).

47. Younes, M. Measurement and testing of respiratory drive, (Dekker, 1981).

48. Blain, G.M., Smith, C.A., Henderson, K.S. & Dempsey, J.A. Contribution of the carotid body chemoreceptors to eupneic ventilation in the intact, unanesthetized dog. Journal of applied physiology 106, 1564–1573 (2009).

49. Paton, J.F., et al. The carotid body as a therapeutic target for the treatment of sympathetically mediated diseases. Hypertension 61, 5–13 (2013).

50. Dejours, P. Chemoreflexes in Breathing. Physiological Reviews 42, 335–358 (1962).

51. Giannoni, A., et al. Combined Increased Chemosensitivity to Hypoxia and Hypercapnia as a Prognosticator in Heart Failure. Journal of the American College of Cardiology 53, 1975–1980 (2009).

52. Smith, C.A., Harms, C.A., Henderson, K.S. & Dempsey, J.A. Ventilatory effects of specific carotid body hypocapnia and hypoxia in awake dogs. Journal of applied physiology 82, 791–798 (1997).

53. Tipton, M.J., Harper, A., Paton, J.F.R. & Costello, J.T. The human ventilatory response to stress: rate or depth? J Physiol 595, 5729–5752 (2017).

54. Gitt, A.K., et al. Exercise anaerobic threshold and ventilatory efficiency identify heart failure patients for high risk of early death. Circulation 106, 3079–3084 (2002).

55. Gluck, S.L. Acid-base. Lancet 352, 474–479 (1998).

56. Skatrud, J.B. & Dempsey, J.A. Interaction of sleep state and chemical stimuli in sustaining rhythmic ventilation. J Appl Physiol Respir Environ Exerc Physiol 55, 813–822 (1983).

57. Xie, A., Skatrud, J.B., Puleo, D.S., Rahko, P.S. & Dempsey, J.A. Apnea-hypopnea threshold for CO2 in patients with congestive heart failure. Am J Respir Crit Care Med 165, 1245–1250 (2002).

58. Taverne, J., et al. High incidence of hyperventilation syndrome after COVID-19. J Thorac Dis 13, 3918–3922 (2021).

59. Ding, Y., Li, Y.L. & Schultz, H.D. Role of blood flow in carotid body chemoreflex function in heart failure. J Physiol 589, 245–258 (2011).

60. Netland, J., Meyerholz, D.K., Moore, S., Cassell, M. & Perlman, S. Severe acute respiratory syndrome coronavirus infection causes neuronal death in the absence of encephalitis in mice transgenic for human ACE2. J Virol 82, 7264–7275 (2008).

61. Guntur, V.P., et al. Signatures of Mitochondrial Dysfunction and Impaired Fatty Acid Metabolism in Plasma of Patients with Post-Acute Sequelae of COVID-19 (PASC). Metabolites 12(2022).

62. Holmes, A.P., Turner, P.J., Buckler, K.J. & Kumar, P. Moderate inhibition of mitochondrial function augments carotid body hypoxic sensitivity. Pflugers Arch 468, 143–155 (2016).

63. Lataro, R.M., et al. P2X3 receptor antagonism attenuates the progression of heart failure. Nat Commun 14, 1725 (2023).

64. McGarvey, L.P., et al. Efficacy and safety of gefapixant, a P2X(3) receptor antagonist, in refractory chronic cough and unexplained chronic cough (COUGH-1 and COUGH-2): results from two double-blind, randomised, parallel-group, placebo-controlled, phase 3 trials. Lancet 399, 909–923 (2022).

65. Excellence, N.I.f.H.a.C. COVID-19 rapid guideline: managing the long-term effects of COVID-19. NG188(2020).

66. Williams, B., et al. 2018 Practice guidelines for the management of arterial hypertension of the European Society of Hypertension (ESH) and the European Society of Cardiology (ESC). Blood Press 27, 314-340 (2018).

67. Graham, B.L., et al. Standardization of Spirometry 2019 Update. An Official American Thoracic Society and European Respiratory Society Technical Statement. Am J Respir Crit Care Med 200, e70–e88 (2019).

68. Quanjer, P.H., et al. Multi-ethnic reference values for spirometry for the 3-95-yr age range: the global lung function 2012 equations. Eur Respir J 40, 1324–1343 (2012).

69. ATS/ACCP Statement on cardiopulmonary exercise testing. Am J Respir Crit Care Med 167, 211-277. (2003).

70. Arena, R., et al. Development of a ventilatory classification system in patients with heart failure. Circulation 115, 2410–2417 (2007).

71. Chua, T.P. & Coats, A.J. The reproducibility and comparability of tests of the peripheral chemoreflex: comparing the transient hypoxic ventilatory drive test and the single-breath carbon dioxide response test in healthy subjects. Eur J Clin Invest 25, 887–892 (1995).

