## Supplementary material for "Carotid body dysregulation contributes to the enigma of long COVID"

### *Exclusion criteria*

*All participants*

- Body mass index ⩾35 kg/$m^2$
- Diagnosed with severe asthma or daily use of inhaler, and/or treatment with oral steroids
- Pregnancy/breastfeeding women
- Ongoing requirement of oxygen therapy
- Taking antihypertensive, nitrate, steroid or immunosuppressant medication or medication
- Major illness e.g., cancer, inflammatory disease (including vasculitis) or receiving palliative care
- History of organ transplantation or are candidates for organ transplantation at the time of screening
- History of Chronic Fatigue Syndrome prior to COVID-19 infection
- Diagnosed cardiovascular disease (including current non-benign arrhythmia, chronic heart failure)
- History of major psychiatric disorder including bipolar disorders, schizophrenia, schizoaffective disorder, major depression.
- Diagnosis of structural lung disease (such as COPD or pulmonary fibrosis)
- Diagnosed renal disease
- Congenital or acquired neurological conditions (including dementia), language disorders, repeated or chronic pain conditions (excluding menstrual pain and minor sporadic headaches)
- Diabetes Mellitus
- Symptoms of febrile illness 2 weeks before experiment
- Excessive alcohol consumption (>28 units/week) or use of illicit drugs
- History of smoking within 2 months
- Surgery under general anaesthesia within 3 months
- History of stroke
- Coronary revascularisation

- Haemodialysis or peritoneal dialysis
- Participating in another study for an investigational medicinal product

### Supplementary results

*Sit-to-stand test*

SBP did not change over time (main effect of time; P=0.2254) but DBP increased over time from sitting to standing (main effect of time; P=0.0484), where DBP increased from rest (78 ± mmHg) to 1 min (84 ± 10 mmHg, P=0.0054), 2 min (85 ± 12 mmHg, P=0.0026) and 3 mins (84 ± 13 mmHg, P=0.0242) of standing. There was no time*group interaction effect for SBP or DBP indicating that both groups responded to sit-to-stand in a similar way. HR increased from sitting to standing (main effect of time, P<0.0001). Unexpectedly, the increase in HR from sit-to-stand was greater in the control group versus the long COVID group (main Time*Group effect; P=0.0258, supplementary figure 2), where the control group had an increase of 13 ± 9 beats/min at 2 mins of standing versus 4 ± 6 beats/min in the long COVID group (P=0.0429). These data indicate that the prevalence of orthostatic intolerance is similar to controls amongst this cohort of long COVID participants.

### Supplementary Tables

**Supplementary table 1:** Medications prescribed to participants in the control and long COVID group. NB: none of these medications were taken on the study visits. Importantly, ivabradine was stopped 48 hours prior to study visits.

| | Controls (n=14) | Long COVID (n=14) |
|---|---|---|
| Ivabradine (n) | 0 | 4 |
| Statins (n) | 1 | 1 |
| Selective serotonin reuptake inhibitors (n) | 2 | 5 |
| Tamsulosin (prostate hyperplasia, n) | 1 | 1 |
| Proton pump inhibitors (n) | 1 | 3 |
| Pain medication (prescribed for myalgia post COVID; pregabalin, n) | 0 | 1 |
| Bivaracetam (epilepsy, n) | 1 | 0 |

**Supplementary table 2**: Blood pressure and heart rate changes during the sit-to-stand test. Data are change from rest.

| | Controls | Long COVID | P-value (mixed-effects ANOVA) |
|---|---|---|---|
| **SBP (mmHg)** | | | |
| **Δ 1 min** | 1.2 ± 10.7 | 6.1 ± 7.9 | Time: P=0.5204 |
| **Δ 2 min** | 1.1 ± 8.7 | 1.9 ± 8.4 | Group: P=0.2836 |
| **Δ 3 min** | -1.6 ± 7.3 | 3.1 ± 9.8 | Time*Group: P=0.7346 |
| **DBP (mmHg)** | | | |
| **Δ 1 min** | 6.9 ± 7.7 | 5.4 ± 7.2 | Time: P=0.1995 |
| **Δ 2 min** | 5.6 ± 9.1 | 6.5 ± 5.8 | Group: P=0.6524 |
| **Δ 3 min** | 2.4 ± 8.8* | 7.3 ± 7.3 | Time*Group: P=0.0012 |
| **Heart rate (beats/min)** | | | |
| **Δ 1 min** | 10 ± 13 | 3 ± 6 | Time: P=0.1631 |
| **Δ 2 min** | 13 ± 9 | 4 ± 6 | Group: P=0.0698 |
| **Δ 3 min** | 10 ± 9 | 6 ± 7 | Time*Group: P=0.1248 |

Mean ± standard deviation. Mixed model ANOVA showed no group or time effects for systolic blood pressure (SBP) or heart rate. There was an interaction effect for DBP where the change in DBP was smaller after 3 mins vs. the change at 1 min (*P=0.0035).

### Supplementary figures

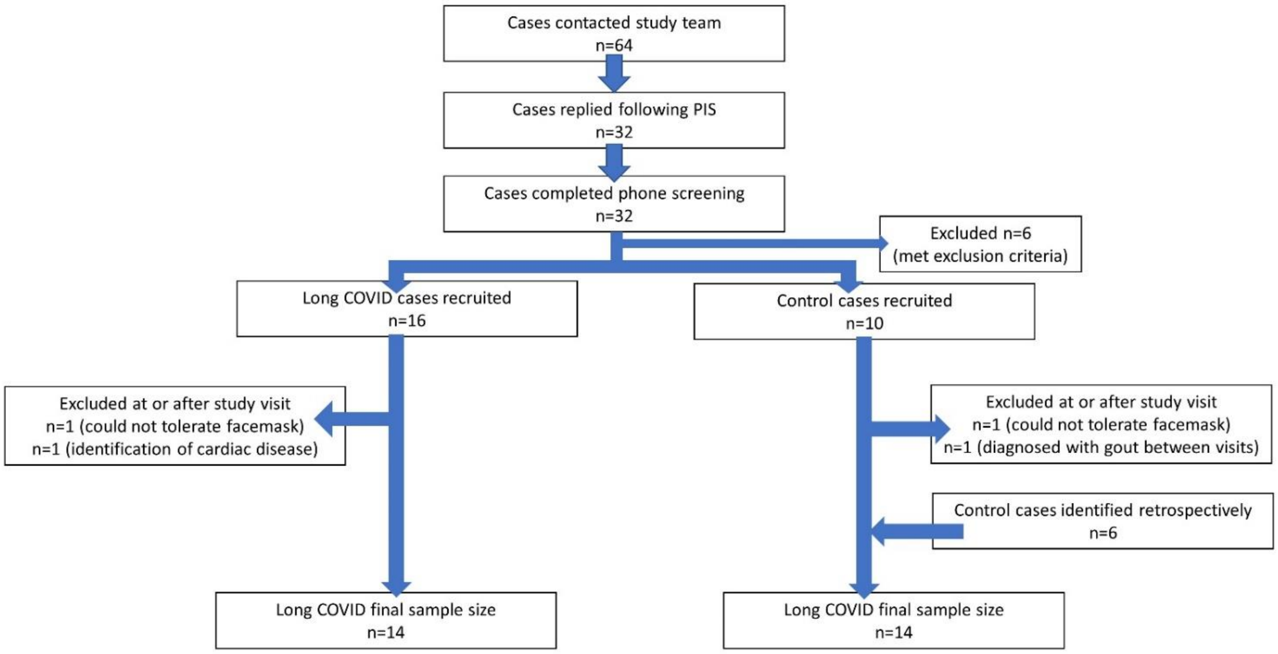


**Supplementary Figure 1:** Flow chart showing recruitment and excluded cases. Following the participant information sheet (PIS) mailout only 50% of individuals interested replied and completed phone screening.

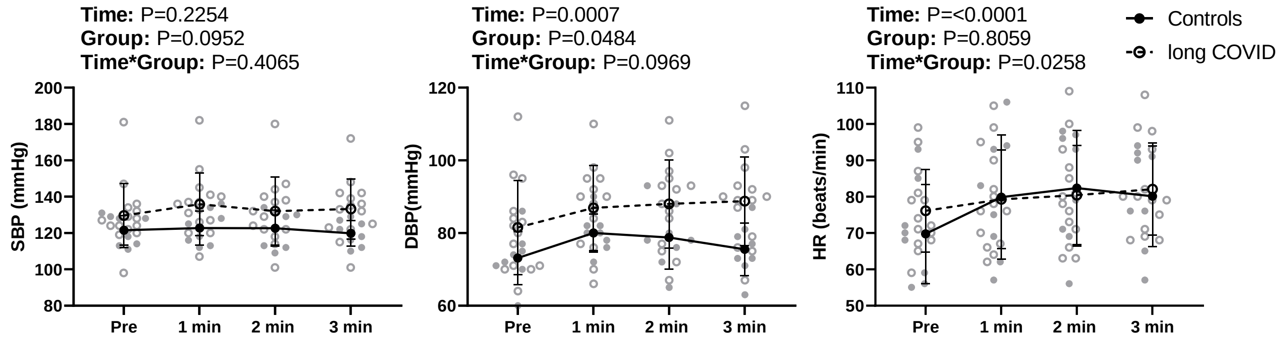


**Supplementary Figure 2:** Sit-to-stand blood pressures and heart rate response in the control and long COVID groups. There were no differences in the BP response to standing between the groups, however, the control group had a greater increase in HR during standing versus the long COVID group. One control and one long COVID participants had HR increases above 100 beats/min. SBP; systolic blood pressure, DBP; diastolic BP and HR; heart rate. Data are mean ± standard deviation.

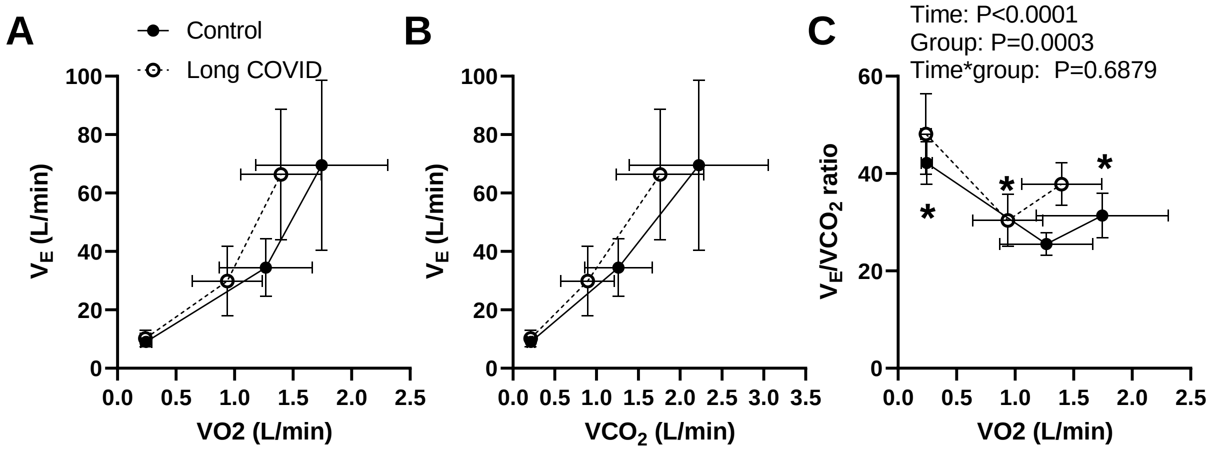


**Supplementary Figure 3:** Ventilation and heart rate during CPET plotted at 3 timepoints: rest, anaerobic threshold, and peak exercise. Panels A and B show the minute ventilation (VE) plotted against the volume of oxygen consumed ($VO_2$) and the volume of $CO_2$ expired ($VCO_2$), respectively. Panel C shows the $V_E/VCO_2$ ratio versus the $VO_2$. The mixed model ANOVA shows that $V_E/VCO_2$ ratio was higher at rest, anaerobic threshold, and peak exercise in the long COVID group. * indicates P<0.05. Rest; P= 0.0031, anaerobic threshold; P=0.0477 and peak exercise; P=0.0051 (Bonferroni's multiple comparison test). Mean ± standard deviation.

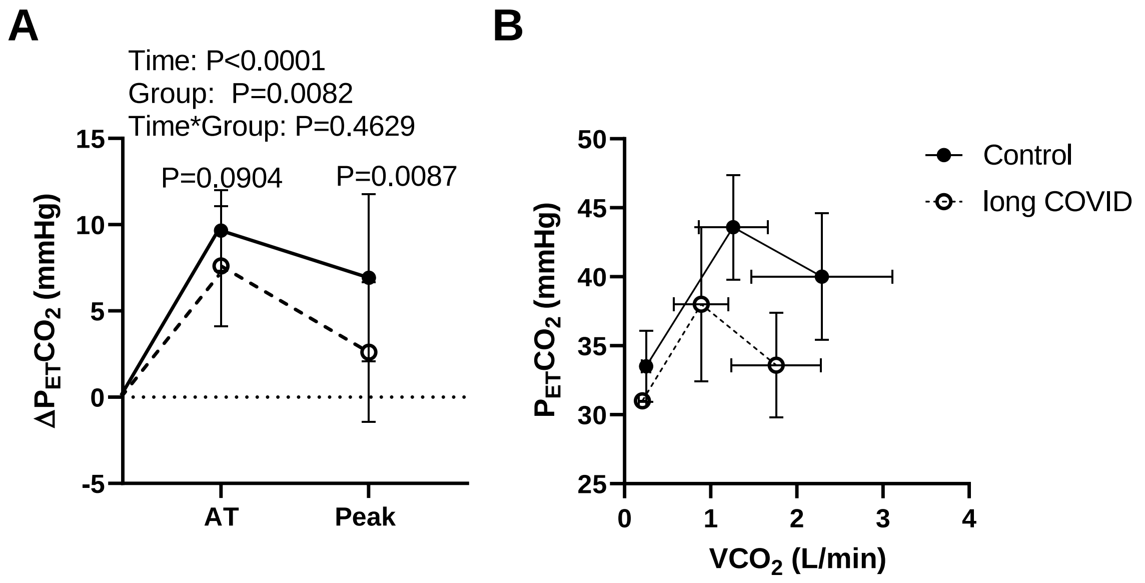


**Supplementary figure 4: A)** Absolute change in partial pressure of end tidal $CO_2$ ($P_{ET}CO_2$) from rest to anaerobic threshold (AT) and peak exercise in the control and the long COVID groups. The long COVID group had a similar increase in $P_{ET}CO_2$ at AT and peak exercise versus the control group. **B)** The $P_{ET}CO_2$ versus the $VCO_2$ plotted at three timepoints from left to right; at rest, at AT and peak exercise in the controls and long COVID group. Data are mean ± standard deviation.

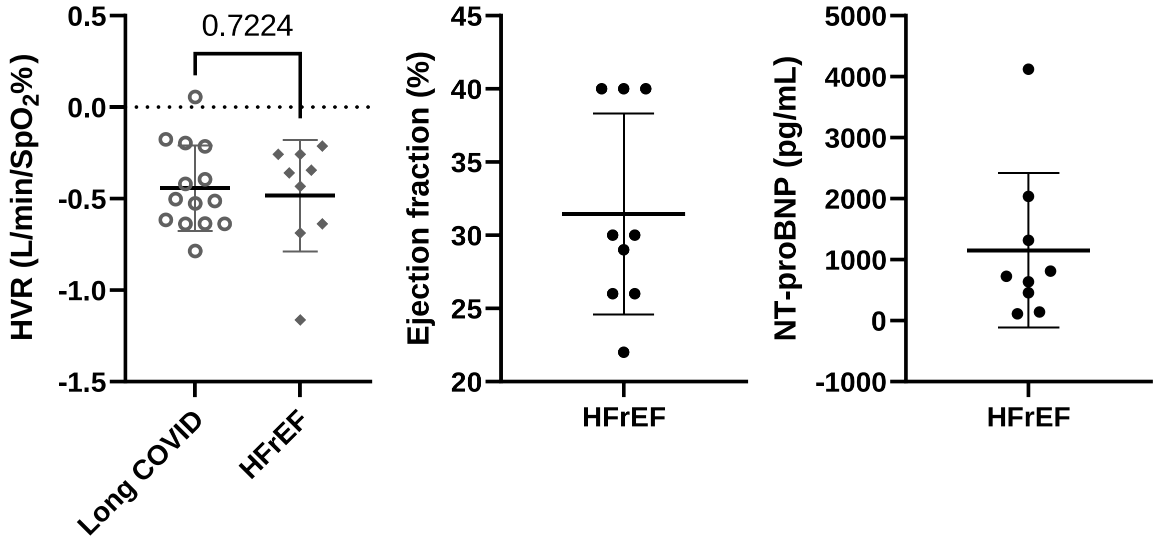


**Supplementary Figure 5:** The hypoxic ventilatory response in the long COVID participants (n=14) versus a group of participants with heart failure reduced ejection fraction (n=9) measured using the same methods, equipment, location, and study team. The heart failure with reduced ejection fraction (HFrEF) participants are taken from our study comparing carotid chemoreflex function in HFrEF versus heart failure with preserved ejection fraction. The NHS research ethics committee approval number is 18/SW/0241. The HVR was -0.44±0.23 L/min/$SpO_2$% versus -0.48±0.30 L/min/$SpO_2$%. Age; 69±11 years, body mass index; 28.7 ± 5.8 kg/$m^2$. Panels B and C show the ejection fraction and NT-proBNP in the HFrEF participants. All participants were prescribed treatment for their heart failure (beta-blockers; n=8, angiotensin converting enzyme inhibitors or angiotensin receptor blocker; n=3, sacubitril with valsartan; n=4, aldosterone antagonist; n=6, ivabradine; n=1) which could impact the hypoxic ventilatory response.
